# Tele-Rapid Response Team (Tele-RRT): Implementation and outcomes of Safety Network System. Before and after cohort study

**DOI:** 10.1101/2021.12.15.21267828

**Authors:** Ahmed N. Balshi, Mohammed A. Al-Odat, Abdulrahman M. Alharthy, Rayan A. Alshaya, Hanan M. Alenzi, Alhadzia S. Dambung, Huda Mhawish, Saad M. Altamimi, Waleed Th. Aletreby

## Abstract

**Background:** Rapid Response Teams were developed to provide interventions for deteriorating patients. Their activation depends on timely detection of deterioration. Automated calculation of warning signs may lead to early recognition, and improvement of RRT effectiveness.

**Method:** This was a “Before” and “After” study, in the “Before” period ward nurses activated RRT after manually recording vital signs and calculating warning scores. In the “After” period, vital signs and warning calculations were automatically relayed to RRT through a wireless monitoring network.

**Results:** The “After” group had significantly lower incidence and rates of cardiopulmonary resuscitation compared to the “Before” group (2.3 / 1000 inpatient days versus 3.8 / 1000 inpatient days respectively, p = 0.01), the “Before” group had a significantly higher hospital length of stay, and significantly fewer visits by the RRT. In multivariable logistic regression model, being in the “After” group decreases odds of CPR by 30% (OR = 0.7 [95% CI: 0.44 – 0.97]; p = 0.02). There was no difference between groups in unplanned ICU admission or readmission.

**Conclusion:** Automated activation of the RRT resulted in significant reduction of CPR events and rate, reduction of hospital length of stay, and increase in the number of visits by the RRT. There was no difference in unplanned ICU admission or readmission.

## Introduction

Hospitalized patients may experience adverse events (AEs), some are as grim as unplanned intensive care unit (ICU) admission, cardiac arrest, and death (1). AEs occur at rates up to 18% in countries with well-developed healthcare systems (2). As many as half of AEs are preventable (3). Crashing patients in hospitals often experience antecedent deterioration of vital signs for an average duration of six hours (1, 4), if such preceding deteriorations were timely recognized, AEs could be prevented by providing simple interventions to prevent further deterioration (5).

Accordingly, several healthcare authorities advocated the concept of Rapid Response Teams (RRT) or sometimes called Rapid Response Systems (RRS) (6, 7).

Regardless of the structure of RRTs which varies among countries and according to hospitals’ needs (8, 9), the team remains the efferent limb of the process, and must be activated to provide further assessment and interventions (10). To be effective, they are dependent on the afferent limb for activation, which uses certain physiological parameters and predefined thresholds of deterioration to trigger RRT activation, such as the National Early Warning Score (NEWS) or its modified version (MEWS) (11, 12). Since calculation of those scores – and subsequently RRT activation - is usually manually performed, and is dependent on staff attentiveness, frequency of measurement, work load, and willingness to call for help (13), it logically follows that there are inevitable delays in RRT activation, which can eventually undermine its effectiveness (1, 14). In fact, several studies described delayed RRT activation as one of the most important causes of lacking efficacy (2).

An attempt to decrease delays in RRT activation, is by real-time recording of vital signs, and automating the calculation of warning scores, however; those modifications didn’t entirely eliminate the need for human activation of RRT, since the alarming vital signs and RRT activation prompts are relayed to the ward nurses (either on central screens or paging), who need to confirm deterioration, and activate RRT manually (11, 13, 15-18). To overcome this gap, tele-monitoring systems were designed to relay the data of deteriorating patients directly to RRTs, eliminating the human factor in activation. Few studies reported the effectiveness of such systems (16), others focused on the system’s performance such as time to complete vital signs recording, completeness of data, and end-users’ satisfaction (19).

In this study we report the effectiveness and patient centered outcomes of a tele-monitoring system that directly relays patients’ vital signs and warning scores to the RRT. We conducted this study under the hypothesis that implementing the system would decrease AEs, due to early activation of RRT.

## Method

This was a cohort study comparing prospective data after an intervention to retrospective data before the intervention. We carried out the study in the Medical tower of the general hospital of King Saud Medical City (KSMC), Riyadh, Saudi Arabia. KSMC is the largest government hospital in the central region of Saudi Arabia, with an in-patient capacity of 1200 beds (300 beds in the Medical tower). The ICU provides an outreach service in the form of a RRT to the general ward composed of an intensivist, two intensive care nurses, and a respiratory therapist. The RRT covers referrals to the ICU by primary admitting specialties, in addition to 48 hours follow up of any patient discharged from the ICU. As per hospital policy, RRT may be deployed if the patients’ vital signs show objective deterioration based on an early alert system, our alert system utilizes criteria of Modified Early Warning Signs (MEWS), and a patient is eligible for RRT activation with a score ≥ 5 (Table 1). It should be mentioned that the RRT is not the only source of admission to ICU from the ward, in fact the majority of ICU admissions from the ward are through the mobile team, which tends to referrals for critically ill patients who are beyond the scope of the RRT. ICU admissions by the RRT are all considered unplanned.

**Table 1:** Modified Early Warning Score:

| **PHYSIOLOGICAL**<br>**PARAMETERS** | **3** | **2** | **1** | **0** | **1** | **2** | **3** |
| --- | --- | --- | --- | --- | --- | --- | --- |
| **Temperature** |  | < 35.0 | 35.1-36.0 | 36.1-38.0 | 38.1-39.0 | > 39.1 |  |
| **Systolic Blood Pressure** | < 90 | 91-100 | 101-110 | 111-200 |  | > 200 | > 220 |
| **Heart Rate** | < 40 |  | 41-50 | 51-100 | 101-110 | 111-130 | > 131 |
| **Respiratory Rate** | < 8 |  | 9 -11 | 12-20 |  |  |  |
| **Oxygen Saturation** | < 91 | 92-93 | 94-95 | > 96 |  |  |  |
| **Any Supplemental Oxygen** |  | YES |  | NO |  |  |  |
| **Level of Consciousness** |  |  |  | A | V | P | U |

A = Alert, V = Reacts to voice, P = Reacts to pain, U = Unconscious.

### Timeframe and data collection

We included data from eight months (January – August) 2020 as the pre-intervention period “Before” group, while the post-intervention period “After” group included the data of the last four months of 2020 and the first four months of 2021 (September 2020 – April 2021). Recorded data included age, gender, whether the patient was a new referral to RRT or being followed for 48 hours after ICU discharge, the main diagnostic medical category (including: Internal medicine, neurology, nephrology, pulmonology, and Hem-Oncology) as we didn’t include surgical patients in the study due to feasibility, MEWS upon RRT activation, and number of RRT activations per patient. We also recorded the length of stay (LOS) of each patient in the ward, reported LOS in this study includes only days in the ward before discharge, ICU admission/readmission, or death. Accordingly, for patients discharged from ICU, LOS doesn’t include the period of stay in the ICU itself. For both groups, we recorded the following outcome variables: ICU admission or readmission, ward CPR and death. Patients transferred to other healthcare facilities were not followed there, and considered as alive discharge.

### Inclusion and exclusion criteria

We included all patients of any age in the medical tower of KSMC, followed by RRT whether they were new activation or follow up after ICU discharge. We excluded the surgical building, patients not followed by the RRT, and patients labelled as “Do Not Resuscitate”.

### Outcomes

The primary outcome measures were occurrence of CPR, and CPR rate. Secondary outcomes included unplanned ICU admission, ICU re-admission for patients discharged from the ICU, and number of RRT activations per patient.

### Intervention

We applied MASIMO SafetyNet ™ (MASIMO, Irvine, CA) to all patients of the “After” group. The system is composed of bed-side sensors that continuously and non-invasively monitor patients’ vital signs, wirelessly sends data to a server capable of simultaneously registering 200 patients, and displays real-time information on a central screen that can display data of up to 40 patients, updated every five minutes. The system also included “Replica”, a mobile application that allows authorized personnel to view patients’ data and receive alarms on smart phones. The software was pre-programmed to alarm visually and audibly when any of the vital signs included in MEWS calculation exceeds its upper or lower limits, as well as when the total MEWS is equal to or more than five based on automated calculation, with only level of consciousness as a manual input by the ward nurse.

MASIMO provided the SafetyNet TM system, however; they were not involved in data collection, analysis, or drafting this study. The study was approved by the local institutional review board with waiver of consent (registration number: H1RI-07-Jan20-02), as the application of the tele-RRT system was considered a performance improvement project. The study follows the ethical principles outlined by the declaration of Helsinki.

### Statistical method

We summarized continuous data as mean ± standard deviation (SD), and discrete data as frequency and percentage (n, %). Compared continuous data by student t test or Wilcoxon rank sum test as appropriate, and discrete data by chi square test or Fisher’s exact test as appropriate. We calculated CPR rate as number of events divided by inpatient days in the ward times 1000 (unit is CPR / 1000 inpatient days).

We fitted a multivariable logistic regression model of predictors of CPR, using backward deletion method to retain significant variables in the model, we evaluated goodness of fit of the model by Hosmer Lemeshow test (well fitted if p > 0.05), and receiver operator characteristic (ROC) curve with area under the curve (AUC). We also explored fulfillment of logistic regression assumptions.

As a sensitivity test for the primary outcome, we fitted a Kaplan Meier curve for survival of both groups, and reported p value of Log-rank test.

All statistical tests were two tailed, considered statistically significant if p value < 0.05 without correction for multiple testing. Commercially available statistical package STATA ® was used in analyses (StataCorp. 2015. Stata Statistical Software: Release 14. College Station, TX: StataCorp LP.).

## Results

The “Before” and “After” groups included data of 2346 and 2151 patients respectively (Figure 1). Notably, DNR rates in both study periods were similar. Table 2 depicts demographic and clinical characteristics of both groups. There were no differences in gender distribution, percentage of patients discharged from ICU, and MEWS at the time of RRT activation.

**Figure 1:**
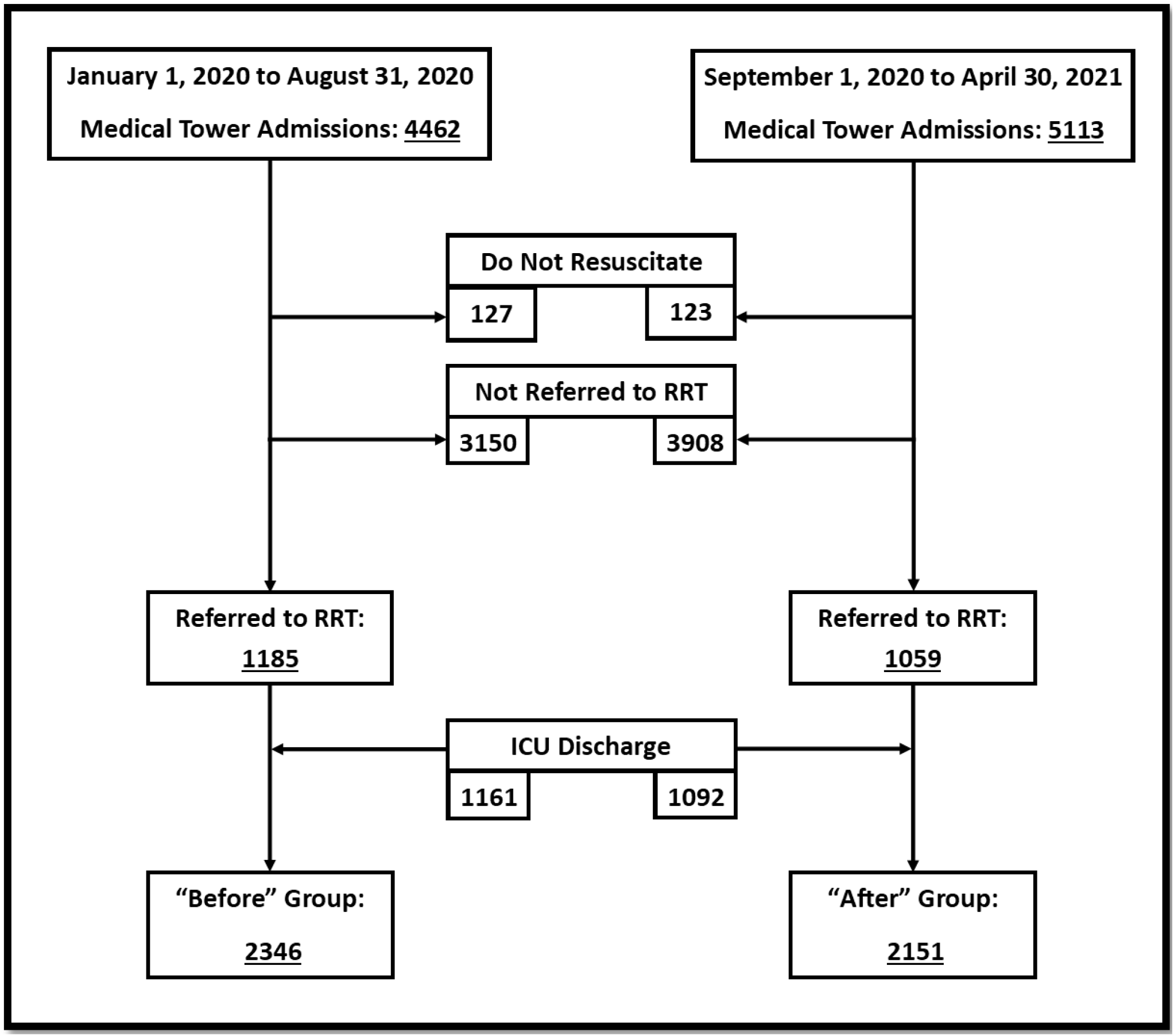
Patients’ enrollment flow diagram:

**Table 2:**
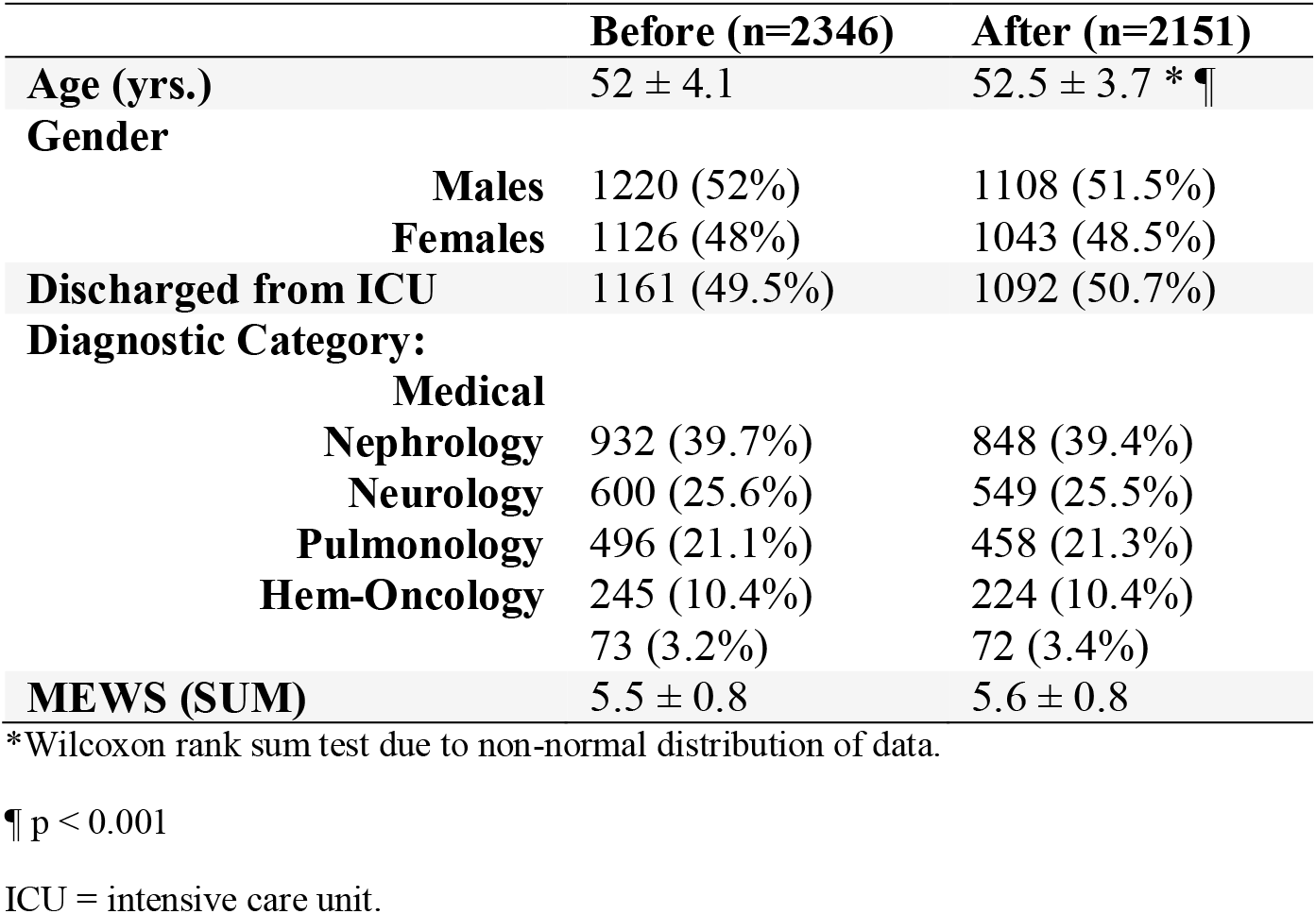
Demographic and clinical characteristics of study groups.

|  | **Before (n=2346)** | **After (n=2151)** |
| --- | --- | --- |
| **Age (yrs.)** | 52 ± 4.1 | 52.5 ± 3.7 * ¶ |
| **Gender** |  |  |
| **Males** | 1220 (52%) | 1108 (51.5%) |
| **Females** | 1126 (48%) | 1043 (48.5%) |
| **Discharged from ICU** | 1161 (49.5%) | 1092 (50.7%) |
| **Diagnostic Category:** |  |  |
| **Medical** |  |  |
| **Nephrology** | 932 (39.7%) | 848 (39.4%) |
| **Neurology** | 600 (25.6%) | 549 (25.5%) |
| **Pulmonology** | 496 (21.1%) | 458 (21.3%) |
| **Hem-Oncology** | 245 (10.4%) | 224 (10.4%) |
|  | 73 (3.2%) | 72 (3.4%) |
| **MEWS (SUM)** | 5.5 ± 0.8 | 5.6 ± 0.8 |

\*Wilcoxon rank sum test due to non-normal distribution of data.
¶ p < 0.001
ICU = intensive care unit.

However, the “After” group had a higher average age compared to “Before” group. In both groups the predominant diagnostic category was Internal medicine, without difference in the distribution of all categories between both groups.

Table 3 shows the primary and secondary outcomes, “Before” group had 78 episodes of CPR, and a total in-patient days of 20,504. This accounts for a CPR incidence of 3.3% and CPR rate of3.8 / 1000 inpatient days (95% CI: 3 – 4.7). Whereas, “After” group had a total of 17,936 inpatient days and 42 CPR episodes, which yielded CPR incidence of 1.95% and CPR rate of 2.3 / 1000 inpatient days (95% CI: 1.7 – 3.2). CPR incidence in the “After” group was significantly lower than “Before” group (p = 0.01). There was no statistical difference in the percentage of unplanned ICU admissions and re-admissions between groups. On the contrary, the “Before” group had a significantly higher average LOS, and significantly fewer visits.

**Table 3:** Primary and secondary outcomes:

|  | Before (n=2346) | After (n=2151) | 95% CI of difference | P value |
| --- | --- | --- | --- | --- |
| CPR (n,%) | 78 (3.3%) | 42 (1.95%) | 0.4% to 2.3% | 0.01 |
| Total in patient days | 20504 | 17936 |  |  |
| CPR Rate: CPR /<br>1000 patient days | 3.8<br>(95% CI: 3 – 4.7) | 2.3<br>(95% CI: 1.7 – 3.2) | 0.3 – 2.6 | 0.01 |
| Unplanned ICU<br>Admission | 28 (1.2%) | 21 (1%) | -0.5% to 0.9 | 0.6 |
| ICU Readmission | 21/1161 (1.8%) | 29/1092 (2.7%) | -0.4% to 2.2% | 0.2 |
| aLOS (days) | 8.7 ± 3.4 | 8.3 ± 3 | 0.2 to 0.6 | < 0.001* |
| Number of visits | 20 ± 7 | 23.7 ± 9.4 | -4.3 to -3.3 | < 0.001* |

\*Wilcoxon rank sum test due to non-normal distribution of data.
CPR = cardiopulmonary resuscitation, ICU = intensive care unit, aLOS = average length of stay, CI = confidence interval.

The logistic regression model of CPR prediction initially included variables of: Age, gender, discharge from ICU, MEWS, number of visits, diagnostic category, and the variable of interest (“Before” or “After” group). Backward deletion method retained three variables in the final model, namely: Age (OR = 1.16 [95% CI: 1.11 – 1.22]; p < 0.001), number of visits (OR = 0.93 [95% CI: 0.91 – 0.96]; p < 0.001), and “After” group (OR = 0.7 [95% CI: 0.44 – 0.97]; p = 0.02).

The model was well fitted (Hosmer Lemeshow p = 0.4), correctly classified 97.3% of the data, and had an AUC of 71.5%. Linear relation between continuous predictors and Log CPR was established by Box-Tidwell test p values > 0.05, and there was no collinearity between continuous predictors (correlation r between Age and number of visits = 0.09) (Figure 2). The significantly lower CPR incidence and rate in the “After” group was supported by a significant p value of Log-rank test (p = 0.01) of the depicted Kaplan Meier curve in the sensitivity analysis (Figure 3).

**Figure 2:**
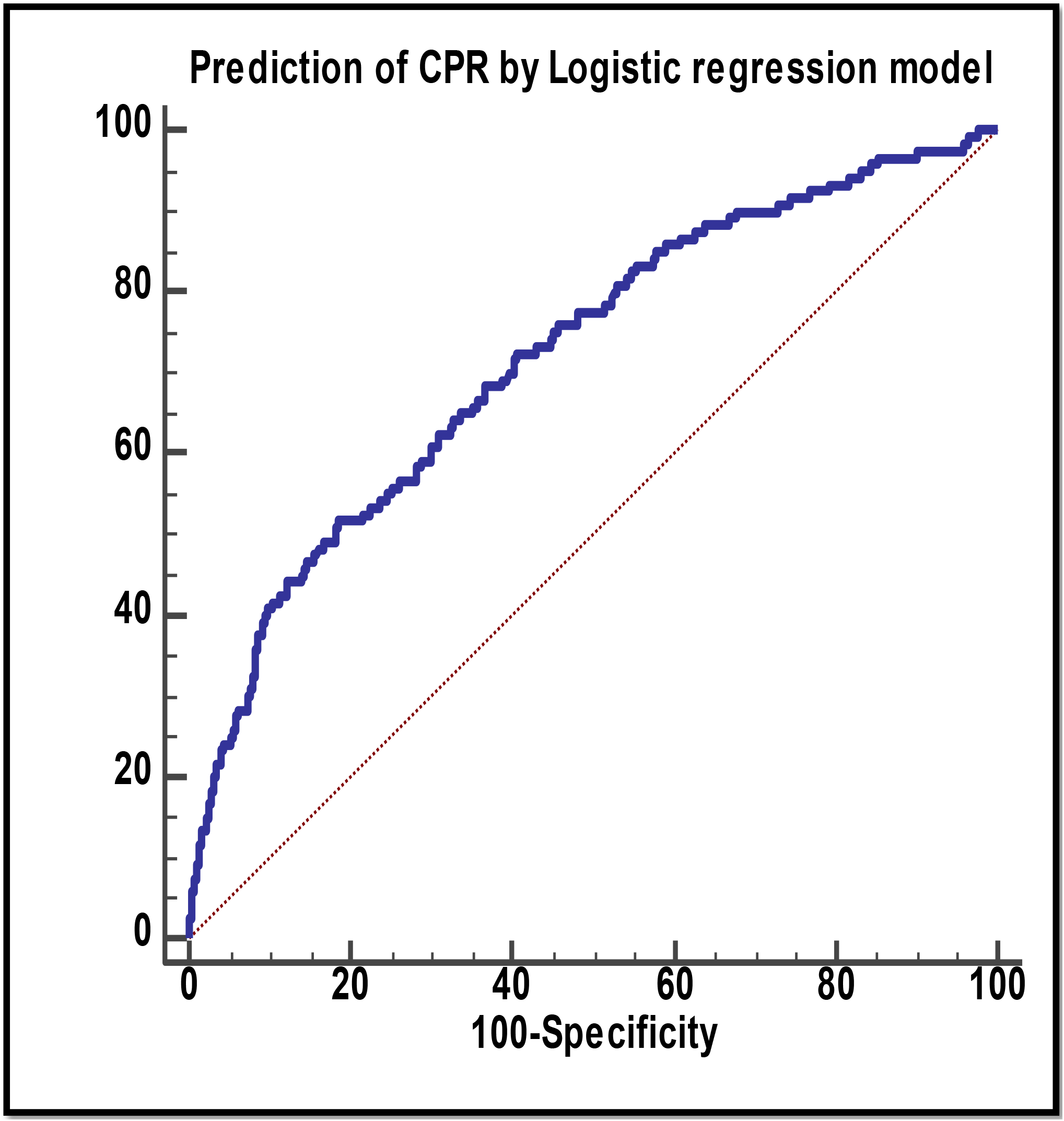
ROC curve of CPR prediction:

**Figure 3:**
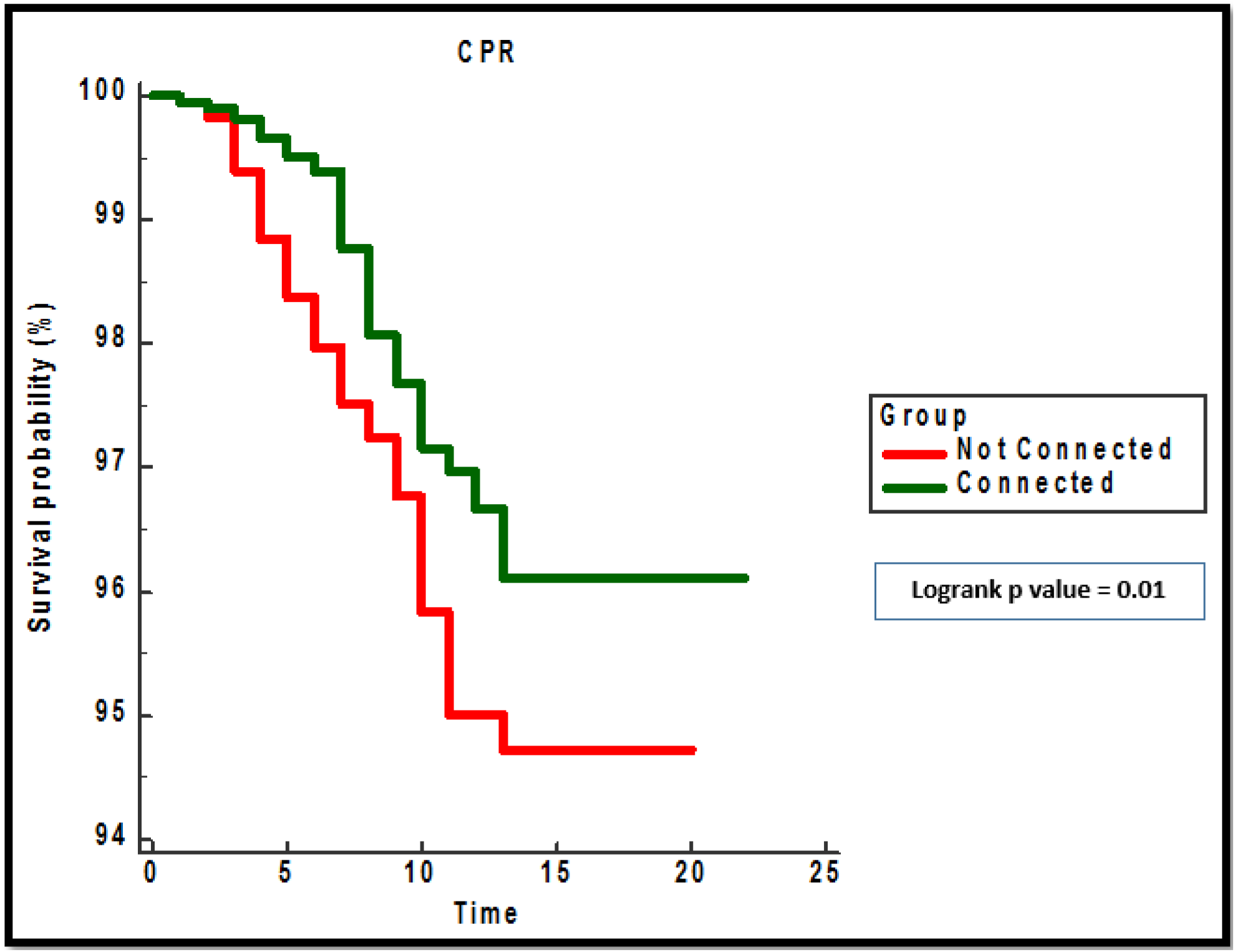
Kaplan Meier Survival curve:

## Discussion

We found in this study that applying MASIMO SafetyNet ™ was associated with lower incidence and rate of CPR, fewer in-patient days, and more visits by RRT. Significant predictors of CPR were Age (OR = 1.16), number of visits (OR = 0.93) and connection to the SafetyNet (OR = 0.7). These results indicate that as age increases by one year odds of CPR increase by 16%, as the number of visits by RRT increase by one odds of CPR decrease by 7%, and being in the “After” group decreases odds of CPR by 30%. These improvements in patients’ outcomes may be attributed to timely recognition of worsening vital signs and deteriorating patients.

Since the introduction of RRTs early in 2000s, several studies questioned their efficacy, showing no improvement of CPR rates, unplanned ICU admission, average LOS, or hospital mortality, among the most famous of which is the MERIT study (20). This may be explained by the dependence of the RRT in its early models on activation by ward staff, who had to assess the patients and calculate warning scores manually, it follows, that if this activation was delayed, the intervention of the RRT would also be delayed, and possibly ineffective. What supports this notion, is that when the systems evolved to automated calculation of warning scores, and electronic prompts of activation, the results of studies – including this study - almost became consistently showing significant improvement particularly in CPR rates, unplanned ICU admission, LOS, and to a lesser extent hospital mortality (13, 16, 17), even when the system still required some manual input such as respiratory rate or level of consciousness. An improvement that was also reflected in systematic reviews (1) despite the inevitable differences between studies (in terms of design – population – RRT structure), thus yielding high heterogeneity percentages which casts a shadow over those benefits.

One of the most commonly reported outcomes of automated RRT systems is the increase in number of visits (16, 21) as it is the case in our study, since there is no human clinical judgement of the patients’ condition, and the activation is entirely based on numerical values. Undoubtedly, this imposes a burden on the team members, and increases their work load. However; from another perspective, frequent visits by the team may be advantageous to the patients, as their condition gets more frequently re-evaluated, and their management regularly updated. We demonstrated this benefit in our study as the reduced odds of CPR as the number of visits increases, in addition to the reduced average hospital LOS, also shown by others (13, 21), possibly as a result of forward progression of management, and prevention of deterioration.

We demonstrated in this study the benefits of applying MASIMO SafetyNet ™ system of automated MEWS calculation and RRT activation. Showing reduced CPR incidence and rate, reducing unplanned ICU admission, and hospital LOS, although coupled with increased number of visits by the RRT. Our study enrolled about 4500 patients, and extended over a period of 16 months. Ranking it among the medium to large studies in terms of size and duration.

Our study suffers numerous limitations. The “Before” and “After” design lacks randomization, which subjects the study to possible bias. We didn’t account for hospital mortality as it customary of similar studies, since the study didn’t include all wards of the hospital. Exclusion of surgical patients is also a limitation, as we can’t generalize our results to that group. The study was conducted during the COVID-19 pandemic, but we didn’t differentiate between COVID-19 positive and negative patients, which may have yielded different results. We only accounted for the diagnostic category when other confounding factors such as comorbidities could have been imbalanced between the study groups. And of course, this was a single center study, reflecting the management in one hospital only, which limits generalizability of its results to the entire region or country.

## Conclusion

Automated activation of the RRT by MASIMO SafetyNet ™ applied to medical wards’ patients resulted in significant reduction of CPR events and rate, reduction of hospital length of stay, and increase in the number of visits by the RRT. There was no difference in unplanned ICU admission or readmission rates.

## Data Availability

All data produced in the present study are available upon reasonable request to the authors

## Acknowledgment

The study team would like to extend their gratitude to all members of the RRT and the medical tower nurses at King Saud Medical City.

We would like to thank Mr. Basil Almuabbadi (Nursing manager of the medical tower), as well as Ms. Bobby Rose Marasigan (project manager of this study).

